# Safety and immunogenicity of a high-dose quadrivalent influenza vaccine administered concomitantly with a third dose of the mRNA-1273 SARS-CoV-2 vaccine in adults ≥ 65 years of age: a Phase II, open-label study

**DOI:** 10.1101/2021.10.29.21265248

**Authors:** Ruvim Izikson, Daniel Brune, Jean-Sébastien Bolduc, Pierre Bourron, Marion Fournier, Tamala Mallett Moore, Aseem Pandey, Lucia Perez, Nessryne Sater, Anju Shrestha, Sophie Wague, Sandrine I Samson

**Affiliations:** Sanofi Pasteur, Swiftwater, Pennsylvania, USA; Accelerated Enrollment Solutions, Peoria, Illinois, USA; Sanofi Pasteur, Marcy l’Etoile, France; Sanofi Pasteur, Lyon, France

**Keywords:** high-dose quadrivalent influenza vaccine, COVID-19 vaccine, booster, coadministration, concomitant administration, influenza vaccination, older adults, public health recommendations

## Abstract

**Background:** Concomitant seasonal influenza vaccination with a COVID-19 vaccine booster could help to minimise potential disruption to the seasonal influenza vaccination campaign and maximise protection against both diseases among individuals at risk of severe disease and hospitalisation. This study assesses the safety and immunogenicity of concomitant administration of high-dose quadrivalent influenza vaccine (QIV-HD) and a mRNA-1273 vaccine booster dose in older adults.

**Methods:** This is an ongoing Phase II, multi-centre, open-label study (NCT04969276). We describe interim results up to 21 days after vaccination (July 2021–August 2021). Adults aged ≥ 65 years living in the community, who were to have received a second mRNA-1273 dose at least five months previously, were randomised (1:1:1) to concomitant QIV-HD and mRNA-1273 vaccination (Coad), QIV-HD alone, or mRNA-1273 vaccine alone. Unsolicited adverse events (AEs) occurring immediately, solicited local and systemic reactions up to day (D)8, and unsolicited AEs, serious AEs (SAEs), AEs of special interest (AESIs) and medically attended AEs (MAAEs) up to D22 were reported. Haemagglutination inhibition (HAI) antibody responses to influenza A/H1N1, A/H3N2, B/Yamagata and B/Victoria strains and SARS CoV-2 binding antibody responses (SARS-CoV-2 Pre-Spike IgG ELISA) were assessed at D1 and D22.

**Findings:** Of 306 participants randomised, 296 were included for analysis (Coad, n=100; QIV-HD, n=92; mRNA-1273, n=104). Reactogenicity profiles were similar between the Coad and mRNA-1273 groups, with lower reactogenicity rates in the QIV-HD group (frequency [95% CIs] of solicited injection site reactions: 86·0% [77·6–92·1], 91·3% [84·2–96·0] and 61·8% [50·9–71·9]; solicited systemic reactions: 80·0% [70·8–87·3], 83·7% [75·1–90·2] and 49·4% [38·7–60·2], respectively). Up to D22, unsolicited AEs were reported for 17·0% and 14·4% participants in the Coad and mRNA-1273 groups, respectively, with a lower rate (10·9%) in the QIV-HD group. Seven MAAEs were reported (Coad, n=3; QIV-HD, n=1; mRNA-1273, n=3). There were no SAEs, AESIs or deaths. HAI antibody geometric mean titres (GMTs) increased from D1 to D22 to similar levels for each influenza strain in the Coad and QIV-HD groups (GMTs [95% confidence interval], range across strains: Coad, 286 [233–352] to 429 [350–525]; QIV-HD, 315 [257–386] to 471 [378–588]). SARS-CoV-2 binding antibody geometric mean concentrations (GMCs) also increased to similar levels in the Coad and mRNA-1273 groups (D22 GMCs [95% confidence interval]: 7634 [6445–9042] and 7904 [6883– 9077], respectively).

**Interpretation:** No safety concerns or immune interference were observed for concomitant administration of QIV-HD with mRNA-1273 booster in adults aged ≥ 65 years, supporting co-administration recommendations.

**Funding:** Sanofi Pasteur

## Introduction

Influenza is an acute viral respiratory disease that affects all age groups, but with a disproportionate disease burden in older adults. Previous global modelling estimates have provided influenza-associated respiratory excess mortality rates of 0·1 to 6·4 per 100 000 individuals for people younger than 65 years, 2·9 to 44·0 per 100 000 individuals for people aged between 65 and 74 years, and 17·9 to 223·5 per 100 000 for people older than 75 years.^1^ Beyond respiratory-related disease, broader non-respiratory consequences of influenza add significant disease burden, including exacerbation of chronic underlying conditions, increased susceptibility to secondary microbial infections, cardiovascular events, and functional decline.^2^

Seasonal influenza vaccination reduces influenza-associated morbidity and mortality in groups at increased risk for complications including older adults.^3,4^ A high-dose influenza vaccine, which contains four times as much haemagglutinin as standard-dose influenza vaccines, was developed to improve protection against influenza in adults aged ≥ 65 years.^5,6^ A meta-analysis of multiple randomised and observational studies conducted over the past 10 years of the high-dose trivalent influenza vaccine (TIV-HD) licensure in the USA, demonstrated that TIV-HD was consistently more effective than standard dose influenza vaccine at reducing influenza disease and associated clinical complications.^7^ To provide broader protection in the context of sustained co-circulation of two B lineages, a quadrivalent formulation of the high-dose vaccine, containing an additional B strain, was licensed by the US Food and Drug Administration (FDA) in November 2019 (QIV-HD; Fluzone^®^ High-Dose Quadrivalent, Sanofi Pasteur).

Coronavirus disease 2019 (COVID-19), caused by severe acute respiratory syndrome coronavirus 2 (SARS-CoV-2), first emerged in humans in December 2019 and spread rapidly to global pandemic status within a few months. The COVID-19 pandemic has had, and continues to have, a devastating impact on public health systems, economies and societies worldwide. Risk factors for severe illness and mortality include age > 75 years, male sex, severe obesity, and active cancer.^8^ The accelerated development and deployment of vaccines against SARS-CoV-2 have helped to limit COVID-19-related severe disease, hospitalisations and deaths in parts of the world where vaccines have been effectively rolled out, with mRNA COVID-19 vaccines among the first to be made available.^9-13^ The mRNA-1273 vaccine (100 µg dose)^14^ has been registered in multiple countries for active two-dose immunisation in individuals aged ≥ 12 years, with some countries also recommending a third 100 µg dose for individuals aged ≥ 18 years with severe immunosuppression.^15,16^ The FDA Vaccines and Related Biological Products Advisory Committee and the US Centers for Disease Control and Prevention’s Advisory Committee on Immunization Practices’ (ACIP) recently expanded eligibility for a 50 µg booster dose to adults with frequent institutional or occupational exposure to SARS-CoV-2 or at high-risk of severe COVID-19, including those aged ≥ 65 years.^17,18^

Reductions in vaccine effectiveness against SARS-CoV-2 infections over time have been observed, which may reflect waning immunity from primary vaccination and/or reduced efficacy against emerging variants.^11-13,19^ Available data on the safety and immunogenicity of third doses of SARS-CoV-2 vaccines mRNA-1273, BNT162b2 and ChAdOx1, support the administration of a booster vaccine dose to enhance protection against COVID-19.^20-25^ Several countries in the Northern Hemisphere (NH) have thus launched a COVID-19 booster vaccine campaign, mostly targeting individuals at risk, including individuals with autoimmune disease and older adults as these groups were among the first to be vaccinated and therefore more likely to have waning immunity.^26^

The public health measures put in place to curb SARS-CoV-2 transmission in 2020 led to significantly reduced influenza activity during the 2020-2021 NH influenza season;^27,28^ the World Health Organization reported less than 0·2% of tested specimens were positive for influenza virus during that period, compared to 17% for the period 2017–2020.^27^ As countries are starting to lift restrictions on everyday life including travel, and with potentially increased susceptibility of individuals to influenza virus due to reduced circulation the previous season, a consequent resurgence of influenza during the upcoming 2021–2022 influenza season is predicted^29,30^

There is a risk of disruption to, or delay of, the seasonal influenza vaccination campaign through prioritised efforts to vaccinate individuals against COVID-19, particularly in older adults, during the same period. To help maintain influenza vaccine uptake and ease pressure on health systems during the coming NH winter season, the World Health Organization and a number of countries have provided guidance on the concomitant administration of influenza and COVID-19 vaccines, with the aim of shortening the vaccination period, reducing the number of visits to healthcare providers, and minimising missed opportunities to immunise against influenza.^31-36^

The current study describes safety and immunogenicity following concomitant administration of QIV-HD and a third (booster) dose of mRNA-1273 vaccine in adults aged ≥ 65 years in the USA.

## Methods

### Study design and participants

This is an ongoing Phase II, multi-centre, open-label, descriptive study (ClinicalTrials.gov: NCT04969276), conducted in six sites in the USA. The study was initiated on 16 July 2021 and has a planned completion date of 15 February 2022, and included: an active phase, from D1 (enrolment, pre-vaccination blood draw and vaccination) through D22 (post-vaccination blood draw), and a six-month safety follow up (D181). Completion of the active phase of the study, reported here, was on 31 August 2021.

Community-dwelling adults aged ≥ 65 years, previously vaccinated with a two-dose primary schedule of the mRNA-1273 vaccine, were eligible for inclusion. The second dose of the primary mRNA-1273 vaccination series was required to have been received at least five months before enrolment in the study. Individuals with immunodeficiency, or who had received immunosuppressive therapy (preceding six months) or long-term systemic corticosteroid therapy, or who had thrombocytopenia or received anticoagulants in the three weeks preceding inclusion, were excluded from the study. Other exclusion criteria included any vaccination in the four weeks preceding the study, or planned receipt of any vaccine between D1 and D28.

The study conduct is in compliance with the International Council for Harmonisation Good Clinical Practice, and ethical principles derived from international guidelines including the Declaration of Helsinki. All participants provided written informed consent.

### Procedures

Participants were randomised in a 1:1:1 ratio, using a permuted block method stratified by site and by age group (< 75 years and ≥ 75 years), to receive concomitant administration of QIV-HD and mRNA-1273 vaccine (Coad group), QIV-HD alone (QIV-HD group), or mRNA-1273 vaccine alone (mRNA-1273 group). Investigators were provided with scratchable randomisation lists by Sanofi Pasteur biostatistics platform. Investigators scratched the list to reveal the randomisation order and the corresponding study group for each participant.

The QIV-HD vaccine (Fluzone^*®*^ High-Dose Quadrivalent, Sanofi Pasteur) used was the 2021–2022 formulation for the NH influenza season. One dose (0·7 mL) contained 60 μg of haemagglutinin of each of the two A strains (A/H1N1, A/H3N2) and two B strains (from the Yamagata and Victoria lineages). The mRNA-1273 vaccine (Moderna) was supplied by the Biomedical Advanced Research and Development Authority, part of the office of the Assistant Secretary for Preparedness and Response at the US Department of Health and Human Services, and was given as a 0·5 mL dose, containing 100 μg of mRNA.

Participants received one or both vaccines by intramuscular injection in the upper arm – one on each side for the concomitant administration group. Participants who were randomised to the mRNA-1273 group were offered the QIV-HD vaccine after completion of the D22 study visit, on a voluntary basis as part of routine medical care and aligned with recommendations of ACIP; no additional data related to this additional vaccination were collected and vaccine receipt was not considered exclusionary.

Haemagglutination inhibition (HAI) antibody responses to influenza A/H1N1, A/H3N2, B/Yamagata and B/Victoria strains were evaluated using a validated HAI assay performed by Global Clinical Immunology, Sanofi Pasteur laboratory (Swiftwater, PA, USA), as described previously.^37^ The endpoint of the assay is the highest serum dilution in which complete inhibition of haemagglutination occurs. The lower limit of quantification was 1:10. Titres below this level were reported as <10 (1/dil).

The SARS-CoV-2 Pre-Spike IgG enzyme-linked immunosorbent assay was a validated assay, performed by Nexelis (Laval, Quebec, Canada) as previously described.^38^ A reference standard was used to quantify antibodies against SARS-CoV-2 Pre-Spike (ELU/mL). The concentration units were converted from ELU/mL to BAU/mL as follows: Result (BAU/mL) = Result (ELU/mL)/7.9815

### Outcomes

Safety endpoints included the number and frequency of: immediate unsolicited systemic adverse events (AEs) and adverse reactions (ARs) occurring within 30 minutes after vaccine injection; solicited injection site and systemic reactions occurring up to 7 days after injection; unsolicited AEs and ARs up to 21 days after injection; and serious AEs (SAEs), AEs of special interest (AESIs), and medically attended AEs (MAAEs) up to six months after injection (see **Appendix** for definitions). Here, we describe safety endpoints through D22. AEs were assessed for intensity and relatedness to study vaccine (see **Appendix** for intensity grading definitions). The AESIs monitored for each vaccine are listed in the **Appendix**.

To describe the HAI antibody response, the following immunogenicity endpoints were reported for each treatment group: HAI geometric mean titres (GMTs) (1/dil) on D1 and D22, D22/D1 titre ratios, detectable HAI titres (≥ 10), and HAI titres ≥ 40, on D1 and D22, and seroconversion rates (titre < 10 on D1 and post-vaccination titre ≥ 40 on D22; or titre ≥ 10 on D1 and a ≥ 4-fold-rise in titre on D22). To describe the SARS-CoV-2 binding antibody response, the following endpoints were reported for each treatment group: geometric mean concentration (GMCs) of anti-S binding IgG on D1 and D22, and D22/D1 ratios; ≥ two-fold-rise and ≥ four-fold-rise in anti-S binding IgG on D22. Additional analyses were carried out to assess the ratios of post-vaccination GMTs or GMCs, between the Coad and single administration groups, for each influenza strain and for the SARS-CoV-2 antigen.

### Statistical methods

All analyses were descriptive. A total of 300 participants (approximately 100 per treatment group) were planned for enrolment. Safety endpoints were described for each study intervention group in the safety analysis set (SafAS; all participants who received at least one dose of the study vaccine). Immunogenicity endpoints were described for each study intervention group in the immunogenicity analysis set (IAS; subset of randomised participants who received at least one dose of study vaccine). All analyses were performed according to the vaccine(s) actually received. For the immunogenicity results, 95% confidence intervals (CIs) for the point estimates were calculated using the normal approximation after log transformation for quantitative data (GMC and geometric mean [GM] of individual ratios) and exact binomial distribution (Clopper-Pearson method, quoted by Newcombe) for single proportions. For the ratios of post-vaccination GMTs or GMCs between Coad and single administration groups, a baseline adjustment was applied using an ANCOVA model of the D22 log_10_ titre/concentration with vaccine group and D01 log_10_ titre as factor and covariate, respectively.

## Results

### Study population

A total of 306 participants were enrolled and randomised, of whom 296 received at least one vaccine dose (Coad, n=100; QIV-HD, n=92; mRNA-1273, n=104). A total of 293 participants completed the active phase (through D22); 11 withdrew (Coad group, n=1 QIV-HD group, n=10) and one (QIV-HD) discontinued due to a protocol deviation (**Supplementary Figure 1**). The higher number of withdrawals in the QIV-HD group was due to participants who had wanted to receive a COVID-19 vaccine discontinuing from the study once they were not randomised to receive mRNA-1273. One participant randomised to the mRNA-1273 group was mistakenly administered both the QIV-HD and mRNA-1273 vaccine and was therefore included in the Coad group for safety and immunogenicity analyses. Overall, in the SafAS, there were more female (166/296 [56·1%]) than male participants, particularly in the mRNA-1273 group (**Table 1**). The mean age of participants was 72 years; 71/296 (24·0%) participants were ≥ 75 years old and the age distribution was balanced between treatment groups. Most participants (282/296 [95·3%]) were white and were non-Hispanic or Latino; and most had received seasonal influenza vaccination the previous season (2020–2021) (**Table 1**). The characteristics of participants in the IAS were comparable to those in the SafAS (data not shown).

**Table 1.**
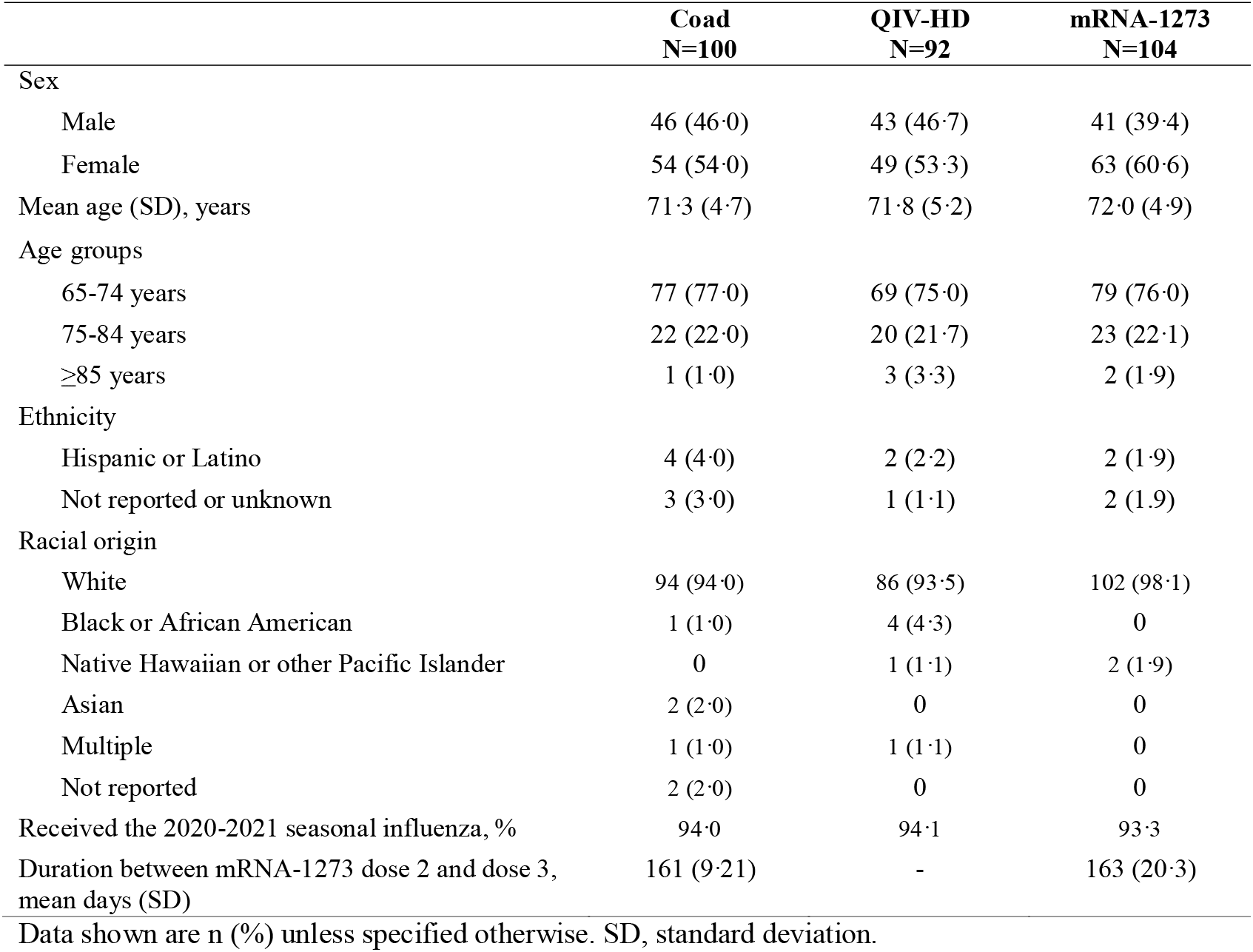
Population characteristics at baseline (Safety analysis set))

### Safety profile

Four immediate unsolicited AEs were reported: three by one participant in the Coad group (anxiety and dizziness, both grade 1 intensity and resolved spontaneously in one day; and grade 3 hypertension, which resolved by D10) and one in a participant in the QIV-HD group (dizziness, grade 1 intensity and assessed by the investigator as related to study vaccination; resolved spontaneously after one day) (**Supplementary Table S1**). Solicited injection-site reactions occurring up to 7 days after mRNA-1273 vaccine injection were reported at similar rate in the Coad group (in the mRNA-1273-injected limb [82·0%; 82/100]) and the mRNA-1273 group (91·3%; 95/104). Those occurring after QIV-HD injection were reported less frequently than after mRNA-1273 vaccine injection, and at similar rates between the Coad group (QIV-HD-injected limb; 61·0%; 61/100) and the QIV-HD group (61·8%; 55/89) (**Supplementary Table S1**). Injection site pain was the most frequently reported solicited injection site reaction in each treatment group; the most frequently reported grade 3 injection site reactions were pain (ranging between 0 and 4·0% per group) and erythema (ranging between 0 and 6·7%) (**Figure 1A**). Solicited systemic reactions were also reported at a similar rate between the Coad and mRNA-1273 groups (80·0% [80/100] and 83·7% [87/104]), and at a lower rate in the QIV-HD group (49·4% [44/89]) (**Supplementary Table 1**). The most frequently reported solicited systemic reaction in each group was fatigue (**Figure 1B**). The most frequently reported grade 3 solicited systemic reactions were malaise, fatigue, and myalgia in the Coad group and malaise, fatigue, headache, and myalgia in the mRNA-1273 group (ranging from 4·8% to 7·0%); only one participant reported a grade 3 reaction (fatigue) in the QIV-HD group.

**Figure 1.**
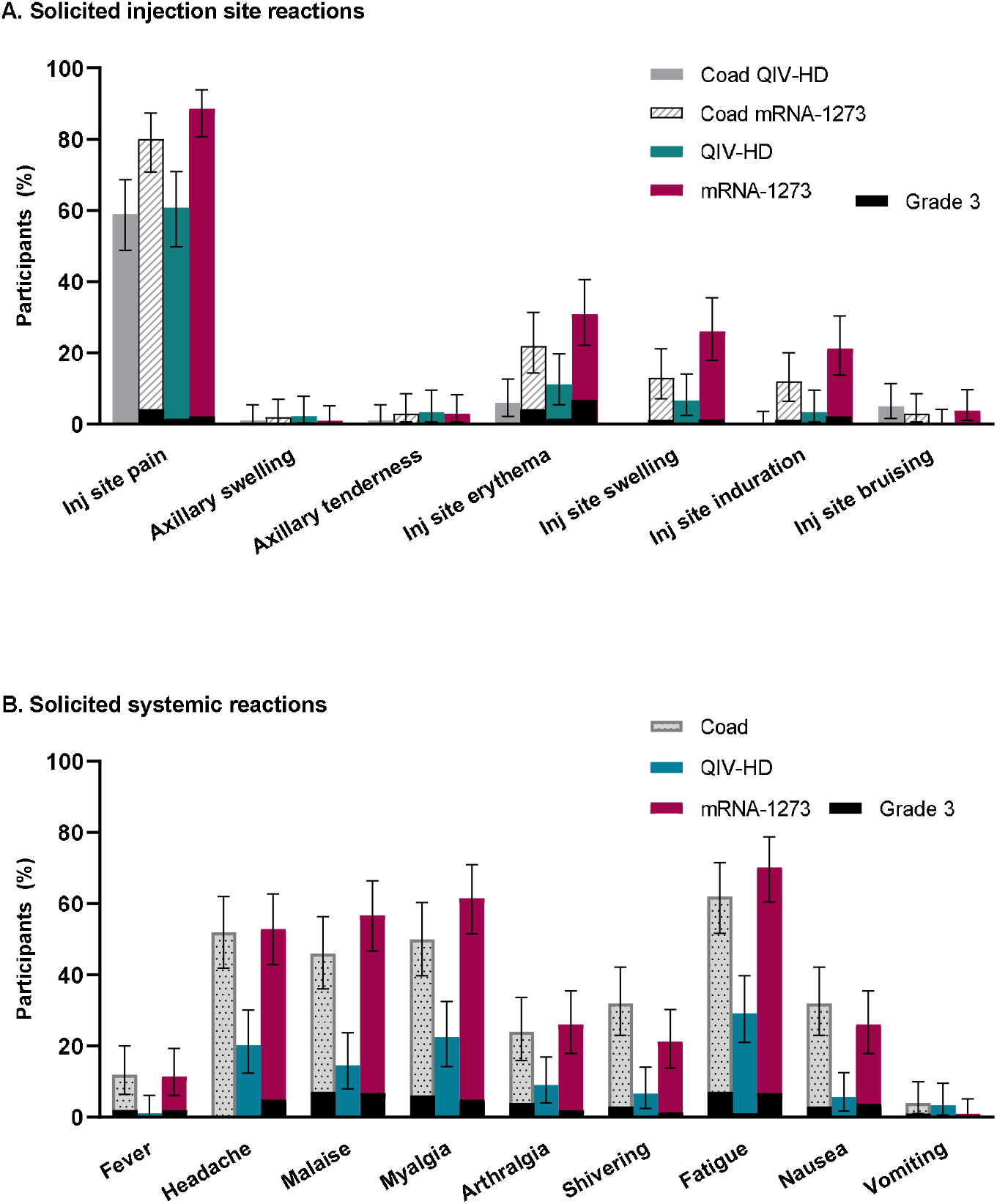
Solicited injection site reactions and systemic reactions occurring up to 7 days post-injection (immunogenicity analysis set) Footnote: Error bars show 95% confidence intervals. Coad QIV-HD, solicited reactions observed in the QIV-HD injected limb of participants in the Coad group. Coad mRNA-1273, solicited reactions observed in the mRNA-1273 injected limb of participants in the Coad group.

Unsolicited AEs through D22 were reported at similar frequencies between the Coad and mRNA-1273 groups (17·0% [17/100] and 14·4% [15/104], respectively), and were reported less frequently in the QIV-HD group (10·9% [10/92]). Two grade 3 unsolicited AEs were reported, one each in the Coad and QIV-HD groups (**Supplementary Table S1**): hypertension (immediate AE) and chemical burns to the eye, respectively. Four participants reported five unsolicited injection site ARs: two participants in the Coad group experienced injection site pruritis (for one participant, in both limbs with onset on D6 and resolving by D8; for the other participant, in the mRNA-1273 injected limb with onset on D2 and resolving by D4); one participant reported injection site warmth after injection of QIV-HD alone, with onset on the day of injection and resolving on D5; and one participant reported injection site pruritis on D3 after injection of mRNA-1273 alone, which resolved on D5. Unsolicited systemic ARs were reported for four participants in the Coad group (n=4 ARs), three participants in the QIV-HD group (n=4 ARs) and six participants in the mRNA-1273 group (n=9 ARs). These included a MAAE of muscle spasms occurring in the left calf of a participant in the mRNA-1273 group, six days post-injection; the spasms were grade 1 intensity and resolved after 2 days with healthcare contact. No grade 3 unsolicited ARs were reported through D22. There were no SAEs, AESIs or deaths reported through D22. Seven participants each reported one MAAE (Coad group, n=3; QIV-HD group, n=1; mRNA-1273 group, n=3).

### Immunogenicity

#### Haemagglutination inhibition antibody responses

For each influenza strain, HAI GMTs were similar across all groups at baseline (D1), and increased post-vaccination at D22 to similar levels in the Coad and QIV-HD groups. Post-vaccination HAI GMTs remained close to baseline levels in the mRNA-1273 group as expected (**Figure 2A**). D22 GMT ratios relative to baseline are presented in **Table 2**. From D1 to D22 the proportion of participants with HAI titres ≥ 40 for each strain increased in the Coad and QIV-HD groups, to similar levels between the two groups (**Figure 3**). Seroconversion rates were similar between the Coad and QIV-HD groups for each strain (**Table 2**).

**Figure 2.**
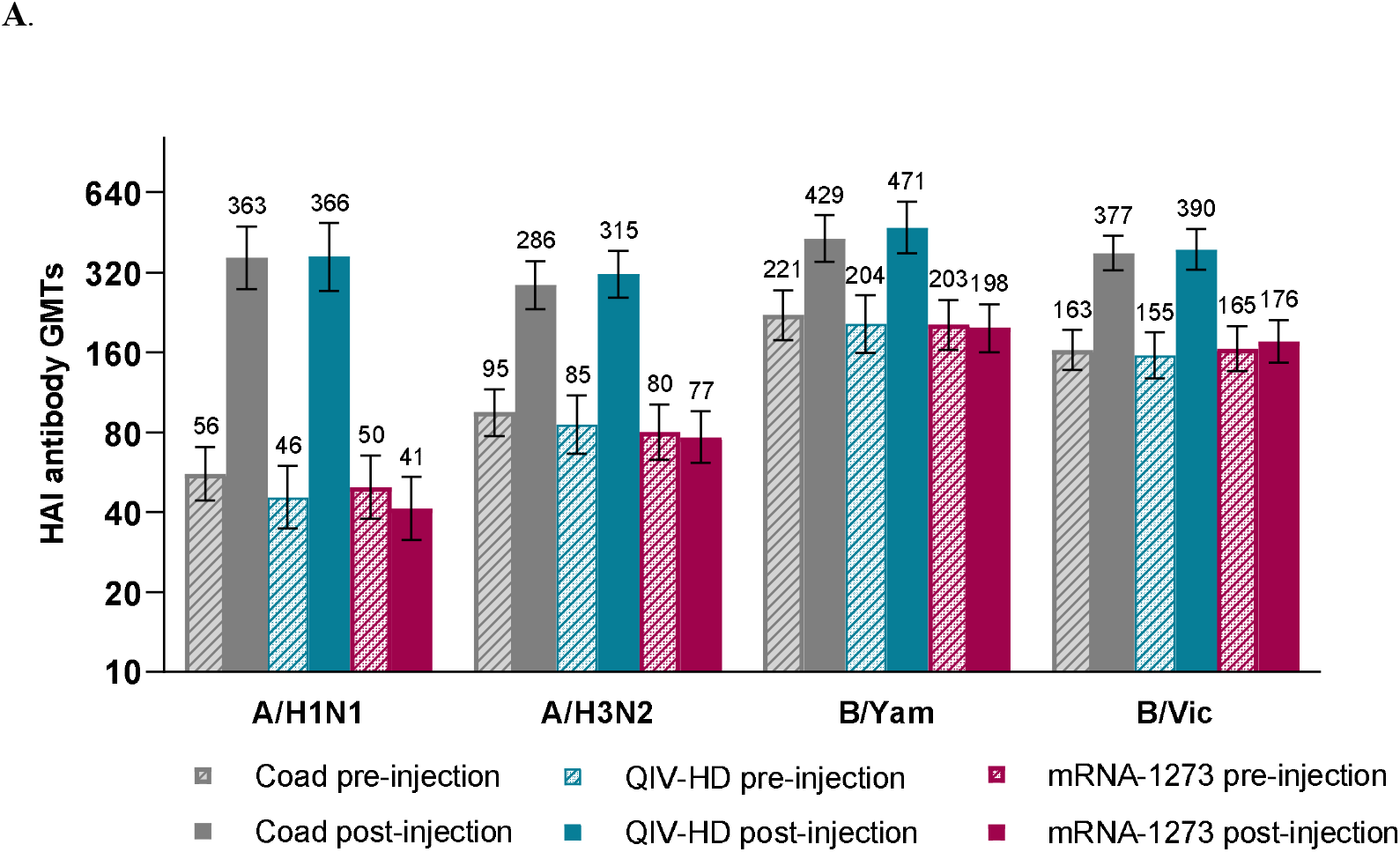
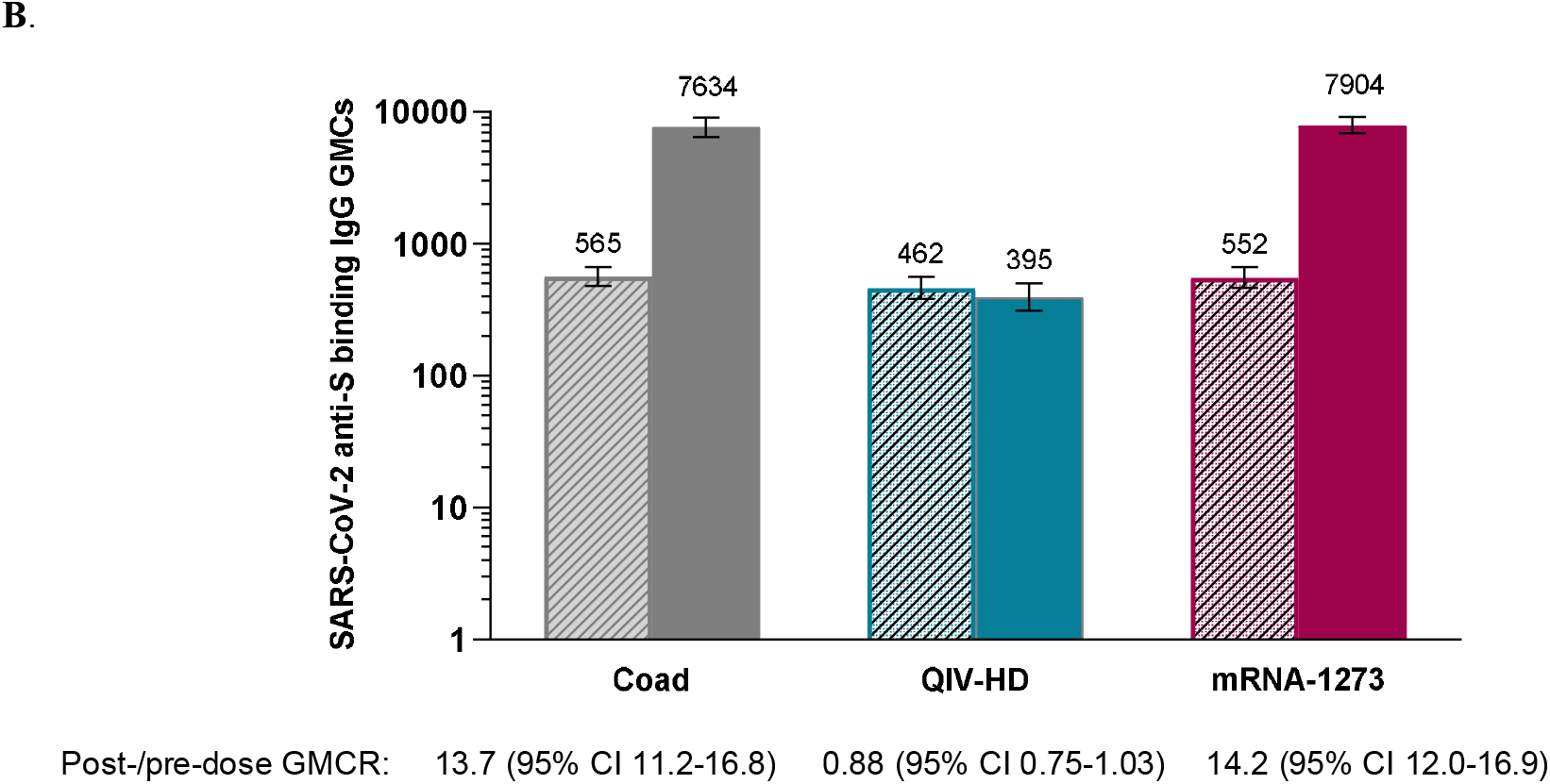
Influenza haemagglutinin inhibition (A) and SARS-CoV-2 anti-S binding IgG (B) antibody responses on Day 1 and Day 22 for each treatment group (immunogenicity analysis set). Footnote: GMC, geometric mean concentration. GMT, geometric mean titre. HAI, haemagglutinin inhibition. Error bars show 95% confidence intervals. Annotations above each bar indicate GMTs (A) or GMCs (B) for each group/time point.

**Table 2.**
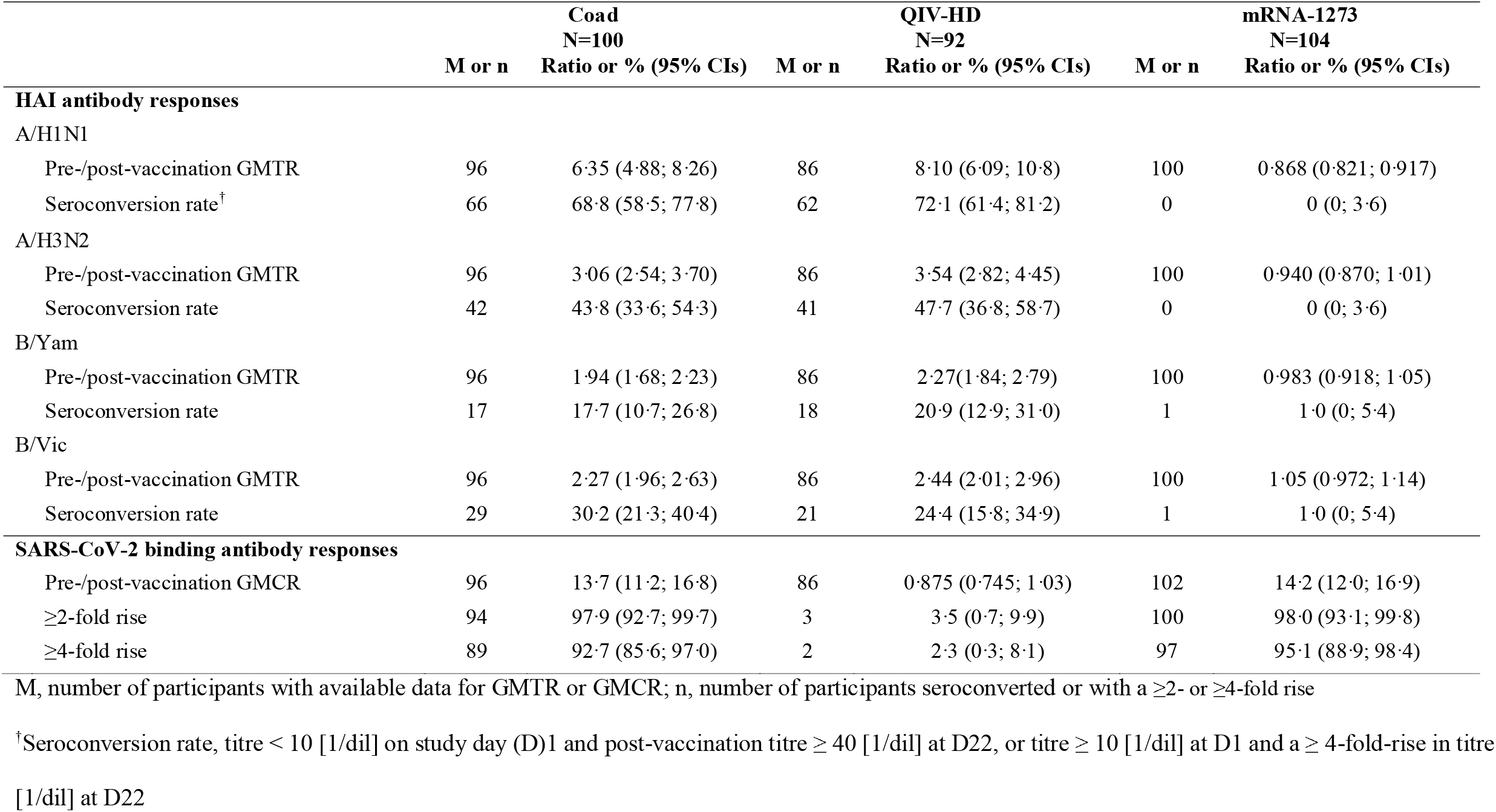
Haemagglutinination inhibition and SARS-CoV-2 anti-S binding antibody responses post-vaccination (D22) for each influenza strain (immunogenicity analysis set)

|  | **Coad**<br>**N=100** |  | **QIV-HD**<br>**N=92** |  | **mRNA-1273**<br>**N=104** |  |
| --- | --- | --- | --- | --- | --- | --- |
|  | **M or n** | **Ratio or % (95% CIs)** | **M or n** | **Ratio or % (95% CIs)** | **M or n** | **Ratio or % (95% CIs)** |
| **HAI antibody responses** |  |  |  |  |  |  |
| **A/H1N1** |  |  |  |  |  |  |
| Pre-/post-vaccination GMTR | 96 | 6.35 (4.88; 8.26) | 86 | 8.10 (6.09; 10.8) | 100 | 0.868 (0.821; 0.917) |
| Seroconversion rate<sup>†</sup> | 66 | 68.8 (58.5; 77.8) | 62 | 72.1 (61.4; 81.2) | 0 | 0 (0; 3.6) |
| **A/H3N2** |  |  |  |  |  |  |
| Pre-/post-vaccination GMTR | 96 | 3.06 (2.54; 3.70) | 86 | 3.54 (2.82; 4.45) | 100 | 0.940 (0.870; 1.01) |
| Seroconversion rate | 42 | 43.8 (33.6; 54.3) | 41 | 47.7 (36.8; 58.7) | 0 | 0 (0; 3.6) |
| **B/Yam** |  |  |  |  |  |  |
| Pre-/post-vaccination GMTR | 96 | 1.94 (1.68; 2.23) | 86 | 2.27 (1.84; 2.79) | 100 | 0.983 (0.918; 1.05) |
| Seroconversion rate | 17 | 17.7 (10.7; 26.8) | 18 | 20.9 (12.9; 31.0) | 1 | 1.0 (0; 5.4) |
| **B/Vic** |  |  |  |  |  |  |
| Pre-/post-vaccination GMTR | 96 | 2.27 (1.96; 2.63) | 86 | 2.44 (2.01; 2.96) | 100 | 1.05 (0.972; 1.14) |
| Seroconversion rate | 29 | 30.2 (21.3; 40.4) | 21 | 24.4 (15.8; 34.9) | 1 | 1.0 (0; 5.4) |
| **SARS-CoV-2 binding antibody responses** |  |  |  |  |  |  |
| Pre-/post-vaccination GMCR | 96 | 13.7 (11.2; 16.8) | 86 | 0.875 (0.745; 1.03) | 102 | 14.2 (12.0; 16.9) |
| ≥2-fold rise | 94 | 97.9 (92.7; 99.7) | 3 | 3.5 (0.7; 9.9) | 100 | 98.0 (93.1; 99.8) |
| ≥4-fold rise | 89 | 92.7 (85.6; 97.0) | 2 | 2.3 (0.3; 8.1) | 97 | 95.1 (88.9; 98.4) |

M, number of participants with available data for GMTR or GMCR; n, number of participants seroconverted or with a ≥2- or ≥4-fold rise
<sup>†</sup>Seroconversion rate, titre < 10 [1/dil] on study day (D)1 and post-vaccination titre ≥ 40 [1/dil] at D22, or titre ≥ 10 [1/dil] at D1 and a ≥ 4-fold-rise in titre [1/dil] at D22

**Figure 3.**
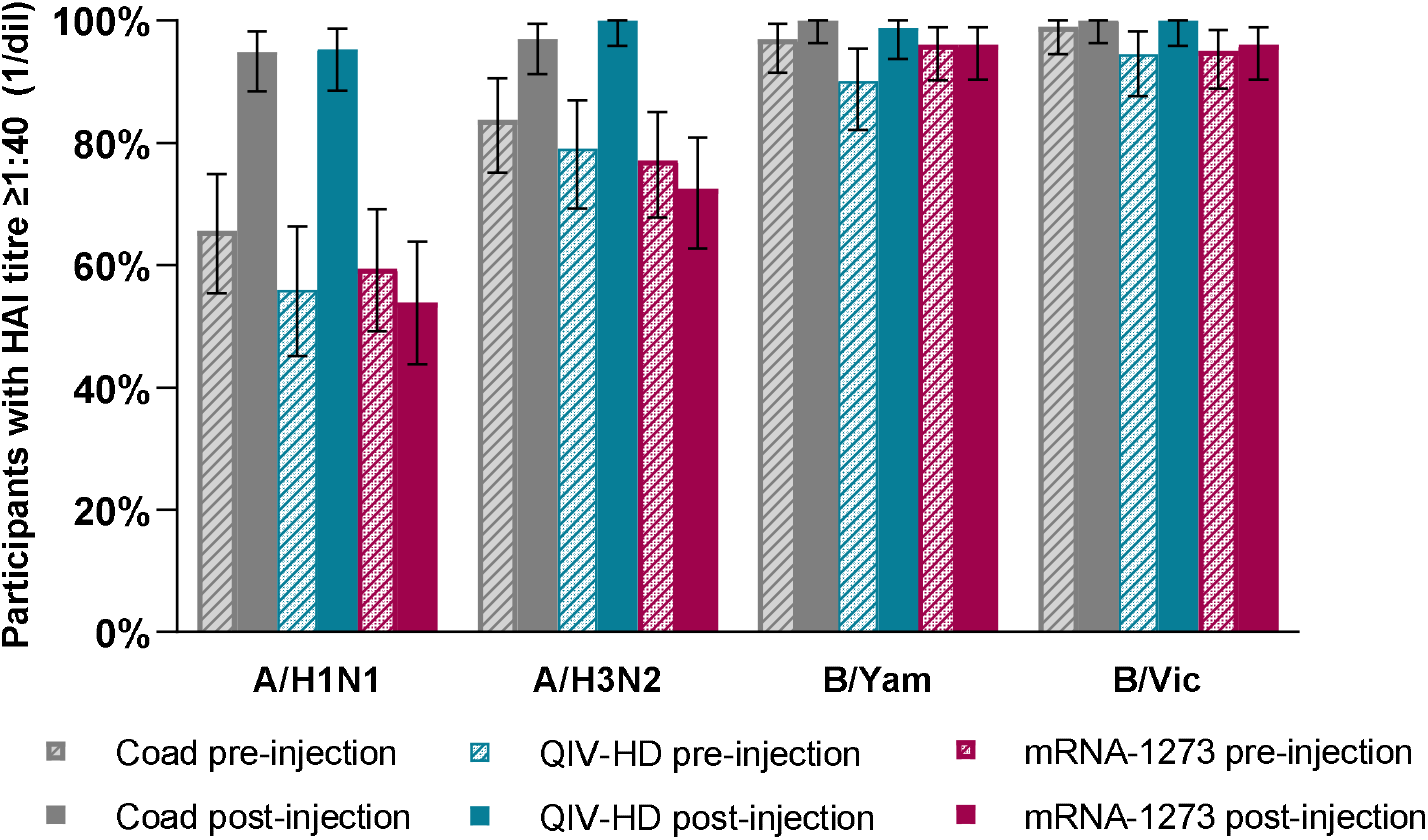
Proportion of participants in each vaccine group with Influenza HAI antibody titres ≥1:40 for each influenza strain pre- (D1) and post-vaccination (D22) (immunogenicity analysis set). Footnote: HAI, haemagglutinin inhibition. Error bars show 95% confidence intervals.

HAI GMT ratios between the Coad and QIV-HD groups at D22 were 0·87 (95% CI 0·61–1·23) for H1N1, 0·89 (95% CI 0·70–1·14) for H3N2, 0·88 (95% CI 0·71–1·09) for B/Yamagata and 0·96 (95% CI 0·79–1·16) for B/Victoria.

#### SARS-Co-V-2 anti-S binding IgG antibody responses

SARS-CoV-2 binding antibody GMCs were similar across all groups at baseline and increased to similar levels in the Coad and mRNA-1273 groups at D22 (7634 and 7904, respectively) (**Figure 2B**). The GMC for participants in the QIV-HD group remained close to baseline levels. At D22, the proportions of participants in the Coad and mRNA-1273 groups with ≥ 2-fold and ≥ 4-fold rises in antibody concentration from baseline were high and similar between groups (**Table 2**).

The SARS-CoV-2 anti-S binding IgG GMC ratio between the Coad and mRNA-1273 groups at D22 was 0·97 (95% CI 0·79–1·19).

## Discussion

In this descriptive interim analysis up to 21 days after vaccination, we did not identify any safety concerns or any evidence of immune interference on influenza HAI or SARS-CoV-2 binding antibody responses after concomitant administration of QIV-HD with a third dose of the mRNA-1273 vaccine (100 µg) in older adults (≥ 65 years) who were immunised with two mRNA-1273 doses approximately five months previously.

We observed similar rates of local reactogenicity in the QIV-HD-injected limb of participants in the Coad group when compared with participants who received QIV-HD alone (any solicited injection site reaction, 61% and 62% respectively) and in the mRNA-1273 injected limb of participants the Coad group when compared with mRNA-1273 vaccine alone (82% and 91.3%, respectively); solicited systemic reactions were reported at similar frequencies in the Coad and mRNA-1273 groups, with lower frequencies observed in participants who received QIV-HD alone. Grade 3 solicited reactions and unsolicited ARs were infrequently reported for all groups and no SAEs, AESIs or deaths were reported. In terms of immunogenicity, comparable HAI antibody responses were observed between the Coad and QIV-HD groups and comparable SARS-CoV-2 binding antibody responses between the Coad and mRNA-1273 groups. The safety and immunogenicity profiles described here for the QIV-HD administered concomitantly with the mRNA-1273 vaccine or administered singly are in-line with previous descriptions of the reactogenicity, safety and immunogenicity of high-dose influenza vaccine in adults ≥ 65 years old^39,40^ or after two 100 µg doses of mRNA-1273 in adults ≥ 55 years old,^41,42^ and are in line with safety descriptions in the QIV-HD and mRNA-1273 vaccine product information, respectively.^43,44^ Our findings of the reactogenicity and immunogenicity of a booster dose of mRNA-1273 vaccine (100 µg) administered concomitantly with QIV-HD or administered alone are also in-line with the results of an open-label phase 2a study demonstrating acceptable safety and immunogenicity of a single booster 50 µg dose of mRNA-1273 in participants who received a two-dose primary series of the COVID-19 vaccine mRNA-1273 approximately 6 months earlier.^21^

A recent study in the UK, involving 679 participants aged ≥18 years (randomised to 12 study arms), showed that the coadministration of a second dose of the SARS-CoV-2 mRNA vaccine BNT162b2 (Pfizer/BioNTech) with one of three seasonal inactivated influenza vaccines had acceptable reactogenicity and tolerability, with no evidence of negative immune interference compared with administration of each vaccine alone.^45^ Based on those preliminary data, the UK government updated their guidance to support concomitant administration of the BNT162b2 vaccine with influenza vaccines (in separate arms)^46^ and encourages concomitant administration of the influenza vaccine with COVID-19 booster vaccination, where practical to do so.^34^

To our knowledge, this study provides the first data supporting concomitant administration of QIV-HD with a COVID-19 booster vaccine dose in older adults. Our data thus provide additional insights that support current public health recommendations to implement seasonal influenza vaccination concomitantly with COVID-19 booster vaccination. This will help to avoid delays in influenza vaccination during the upcoming NH influenza season, particularly among those at increased risk of waning protection from COVID-19 and increased risk of severe illness and hospitalisation due to both COVID-19 and influenza.

This study employs previously validated assays to measure HAI antibody responses^37^ and the SARS-CoV-2 Pre-Spike IgG antibody responses.^38^ High influenza HAI antibody titres were observed pre-vaccination, especially for the influenza B strains. This may be due to the fact that this study population was vaccinated very early in the influenza season (July-August) and that the same B strains were used in the 2020–2021 and 2021–2022 vaccine compositions; moreover, greater than 90% of the subject population had received influenza vaccine in the prior season. Despite the fact that we included approximately 100 participants in each intervention group, this study was not powered for statistical comparisons between study groups. Furthermore, the dose of mRNA-1273 (100 µg) is greater than the dose (50 µg) that has recently been authorised for a booster dose. In the absence of increase in incidence or severity of AEs and ARs and without evidence of interference in the immune response, the results of this study should enable extrapolation to coadministration of lower doses with longer intervals between the second and third dose. Notably, the study population did not sufficiently represent the ethnic and racial diversity of the US population. This was in part due to the very short time window for enrolment of study participants (approximately two weeks), but should be prioritised and improved in future studies.

Our findings suggest that QIV-HD and the mRNA-1273 vaccine can be administered together without evidence of any safety concern or interference in the immune response, thus supporting existing guidance on the implementation of concomitant vaccination campaigns against both influenza disease and COVID-19.

## Data Availability

All data produced in the present study are available upon reasonable request to the authors

## Author Contributions

RI, DB, MF and SIS contributed to the concept or design of the study; DB contributed to data acquisition; and RI, DB, JSB, PB, MF, TMM, AP, LP, NS, AS, SW and SIS were involved in the analysis and/or interpretation of the data. All authors were involved in drafting or critically revising the manuscript, and all authors approved the final version and are accountable for the accuracy and integrity of the manuscript. All authors had full access to all the data in the study and had final responsibility for the decision to submit for publication. At least two authors (RI and PB) have accessed and verified the data.

## Data sharing

Qualified researchers may request access to patient level data and related study documents including the clinical study report, study protocol with any amendments, blank case report form, statistical analysis plan, and dataset specifications. Patient level data will be anonymised and study documents will be redacted to protect the privacy of trial participants. Further details on Sanofi’s data sharing criteria, eligible studies, and process for requesting access can be found at: https://www.clinicalstudydatarequest.com.

## Acknowledgements

The authors would like to thank all participants, investigators and study site personnel who took part in this study. In particular, we would like to thank: Mark H Hutchens, Peter Levins, Isabel Pereira, Thomas Starkey, Patrick Yassini; from Sanofi Pasteur: Iris Depaz, Stephanie Pepin, Kevin Yin, Camille Salamand and Cynthia Tabar; from Moderna; Hamilton Bennett, Julie Vanas, Andrea Sutherland, Deborah Manzo, Amparo Figueroa, Brett Leav; and the US HHS-DoD COVID-19 Countermeasures Acceleration Group (CAG) and BARDA. The authors also acknowledge Juliette Gray of inScience Communications, Springer Healthcare Ltd, London, UK for editorial assistance with the preparation of this manuscript, funded by Sanofi Pasteur, and Isabel Grégoire for editorial assistance and manuscript coordination on behalf of Sanofi Pasteur.

