## Supplementary materials for "Safety and immunogenicity of a high-dose quadrivalent influenza vaccine administered concomitantly with a third dose of the mRNA-1273 SARS-CoV-2 vaccine in adults ≥ 65 years of age: a Phase II, open-label study"

**Appendix**

**Supplementary Methods**

**Assessment of safety - definitions**

*Adverse event (AE)*: An AE is any untoward medical occurrence in a clinical study participant, temporally associated with the use of study intervention, whether or not considered related to the study intervention.

*Adverse Reaction (AR)*: An AR is any noxious and unintended response to a study intervention

related to any dose

*Serious adverse event (SAE):* Any AE that, at any dose, results in death, is life-threatening, requires inpatient hospitalisation or prolongation of existing hospitalisation, results in persistent or significant disability/incapacity, or is another medically important event (not meet any of the above seriousness criteria, but which are considered as serious based on investigator medical judgment).

*Medically Attended AE (MAAE)*: An MAAE is a new onset or a worsening of a condition that prompts the participant or participant’s parent/legally acceptable representative to seek unplanned medical advice at a physician’s office or Emergency Department. Physician contact made over the phone or by email will be considered a physician office visit for the purpose of MAAE collection. This includes medical advice seeking during the study visit or routine medical care. This definition excludes pediatric check-ups, follow-up visits of chronic conditions with an onset prior to entry in the study, and solicited reactions.

*Adverse Event of Special Interest (AESI)*: An AESI (serious or non-serious) is one of scientific and medical concern specific to the Sponsor’s study intervention or program, for which ongoing monitoring and rapid communication by the investigator to the Sponsor can be appropriate. Such an event might warrant further investigation in order to characterize and understand it. Depending on the nature of the event, rapid communication by the study Sponsor to other parties (eg, regulators) might also be warranted.

AESIs were captured as SAEs and included:

| *Fluzone High-Dose Quadrivalent vaccine-related AESIs* | |
| --- | --- |
| - Guillain-Barré Syndrome | |
| - Encephalitis/myelitis (including transverse myelitis) | |
| - Neuritis (including Bell’s palsy, optic neuritis, and brachial neuritis) | |
| - Thrombocytopenia | |
| - Vasculitis | |
| - Convulsions | |
| - Anaphylaxis or other hypersensitivity/allergic reactions | |
| *Moderna COVID-19 Vaccine-related AESIs* | |
| - Anosmia, Ageusia | - Acute liver injury |
| - Subacute thyroiditis | - Dermatologic findings |
| - Acute pancreatitis | - Multisystem inflammatory disorders |
| - Appendicitis | - Thrombocytopenia |
| - Rhabdomyolysis | - Acute aseptic arthritis |
| - Acute respiratory distress syndrome (ARDS) | - New onset of or worsening of neurologic disease |
| - Coagulation disorders | - Anaphylaxis |
| - Acute cardiovascular Injury | - Acute kidney injury |
| Other syndromes:   - Fibromyalgia - Postural Orthostatic Tachycardia Syndrome | - Chronic Fatigue Syndrome - Myasthenia gravis |

**Assessment of safety – intensity grading scale**

For measurable AEs that are part of the list of solicited reactions, the following scale is used:

Grade 1, ≥38·0°C to ≤38·4°C (fever) or ≥25 to ≤50 mm

Grade 2: ≥38·5°C to ≤38·9°C (fever) ≥51 to ≤100 mm

Grade 3: ≥39·0°C (fever) >100 mm.

All other unsolicited AEs will be classified according to the following intensity scale:

• Grade 1: usually transient and may require only minimal treatment or therapeutic intervention. The event does not generally interfere with usual activities of daily living.

• Grade 2: usually alleviated with additional therapeutic intervention. The event causes some interference with activity.

• Grade 3: an event that interrupts usual activities of daily living, or significantly affects clinical status, or may require intensive therapeutic intervention.

.

**Supplementary Table S1. Frequency of unsolicited AEs, 30 min and 21 days post-injection, and solicited injection site and systemic reactions through seven days post-injection**

|  | | **Coad N=100** | | **QIV-HD N=92** | | **mRNA-1273 N=104** | |
| --- | --- | --- | --- | --- | --- | --- | --- |
|  | | **n/M** | **% (95% CI)** | **n/M** | **% (95% CI)** | **n/M** | **% (95% CI)** |
| **Within 30 minutes after vaccine injection** | |  |  |  |  |  |  |
| **Immediate unsolicited AE** | | 1/100 | 1·0 (0; 5·4) | 1/92 | 1·1 (0; 5·9) | 0/104 | 0 (0; 3·5) |
|  | **Immediate unsolicited AR** | 0/100 | 0 (0; 3·6) | 1/92 | 1·1 (0; 5·9) | 0/104 | 0 (0; 3·5) |
| **Within 7 days after vaccine injection** | |  |  |  |  |  |  |
| **Solicited reaction** | | 94/100 | 2·0 (0·2; 7·0) | 65/89 | 73·0 (62·6; 81·9) | 100/104 | 96·2 (90·4; 98·9) |
| **Solicited injection site reaction** | | 86/100 | 1·0 (0; 5·4) | 55/89 | 61·8 (50·9; 71·9) | 95/104 | 91·3 (84·2; 96·0) |
|  | - After injection of QIV-HD | 61/100 | 61·0 (50·7; 70·6) | 55/89 | 61·8 (50·9; 71·9) | - | - |
|  | - After injection of mRNA-1273 | 82/100 | 82·0 (73·1; 89·0) | - | - | 95/104 | 91·3 (84·2; 96·0) |
| **Grade 3 solicited injection site reaction** | | 8/100 | 8.0 (3.5; 15.2) | 2/89 | 2.2 (0.3; 7.9) | 8/104 | 7.7 (3.4; 14.6) |
|  | - After injection of QIV-HD | 0/100 | 0 (0; 3·6) | 2/89 | 2.2 (0.3; 7.9) | - | - |
|  | - After injection of mRNA-1273 | 8/100 | 8.0 (3.5; 15.2) | - | - | 8/104 | 7.7 (3.4; 14.6) |
| **Solicited systemic reaction** | | 80/100 | 80·0 (70·8; 87·3) | 44/89 | 49·4 (38·7; 60·2) | 87/104 | 83·7 (75·1; 90·2) |
| **Grade 3 solicited injection site reaction** | | 13/100 | 13.0 (7.1; 21.2) | 1/89 | 1.1 (0; 6.1) | 14/104 | 13.5 (7.6; 21.6) |
| **Within 21 days after vaccine injection** | |  |  |  |  |  |  |
| **Unsolicited AE** | | 17/100 | 17·0 (10·2; 25·8) | 10/92 | 10·9 (5·3; 19·1) | 15/104 | 14·4 (8·3; 22·7) |
|  | **Grade 3 unsolicited AE** | 1/100 | 1.0 (0; 5.4) | 1/92 | 1.1 (0; 5.9) | 0/104 | 0 (0; 3.5) |
| **Unsolicited AR** | | 6/100 | 6·0 (2·2; 12·6) | 4/92 | 4·3 (1·2; 10·8) | 7/104 | 6·7 (2·7; 13·4) |
|  | **Grade 3 unsolicited AR** | 0/100 | 0 (0; 3.6) | 0/92 | 0 (0; 3.9) | 0/104 | 0 (0; 3.5) |
|  | **Unsolicited injection site AR** | 2/100 | 2·0 (0·2; 7·0) | 1/92 | 1·1 (0; 5·9) | 1/104 | 1·0 (0; 5·2) |
|  | - After injection of QIV-HD | 1/100 | 1·0 (0; 5·4) | 1/92 | 1·1 (0; 5·9) | - | - |
|  | - After injection of mRNA-1273 | 2/100 | 2·0 (0·2; 7·0) | - | - | 1/104 | 1·0 (0; 5·2) |

M, number of participants with available data for the specified endpoint; n, number of participants experiencing the specified endpoint

**Supplementary Figure S1. Participant flow through the study (active phase)**

Footnote: D, study day; IAS, immunogenicity analysis set; n, number of participants meeting specified criteria; SAS, safety analysis set

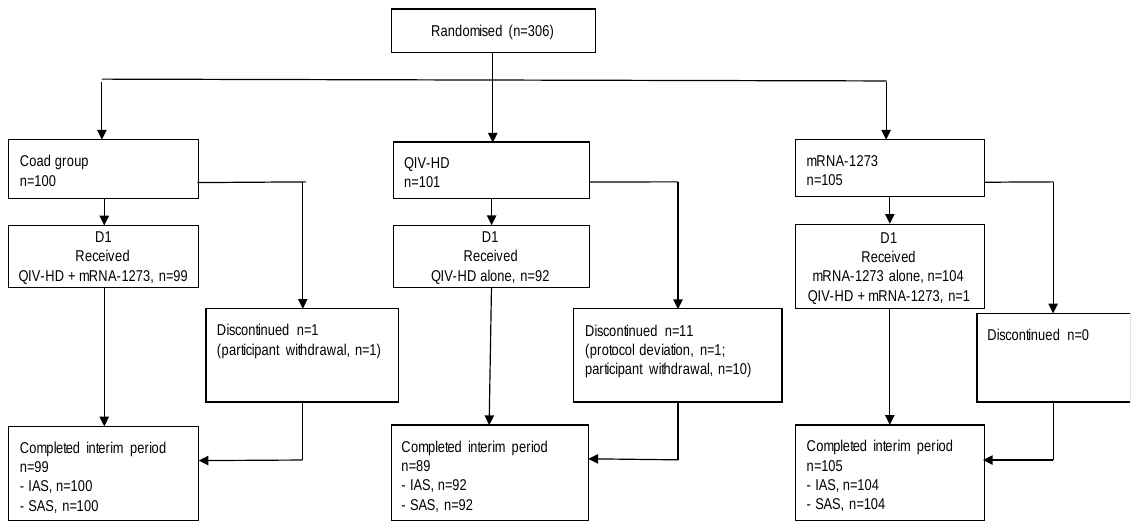
